# The prevalence of chronic kidney disease in Australian primary care: analysis of a national general practice dataset

**DOI:** 10.1101/2023.06.18.23290762

**Authors:** Min Jun, James Wick, Brendon L. Neuen, Sradha Kotwal, Sunil V. Badve, Mark Woodward, John Chalmers, David Peiris, Anthony Rodgers, Kellie Nallaiah, Meg J Jardine, Vlado Perkovic, Martin Gallagher, Paul E. Ronksley

**Author notes:** **Corresponding author:** Min Jun The George Institute for Global Health, Australia Faculty of Medicine and Health, University of New South Wales (UNSW), Sydney, Australia Level 5, 1 King St Newtown NSW 2042 Australia.

## Abstract

**Background:** There remains substantial variation in the reported prevalence of CKD in Australia. Using a large, nationally-representative general practice data source in Australia, we determined the contemporary prevalence and staging of CKD in Australian primary care.

**Methods:** We performed a retrospective, community-based observational study using healthcare data from MedicineInsight, a national general practice data source in Australia. The study included all adults with ≥1 visit to a general practice participating in the MedicineInsight program and ≥1 serum creatinine measurement (with or without a urine albumin-to-creatinine ratio [UACR] measurement) between 1 January 2011 and 31 December 2020; n=2,720,529 patients). The prevalence of CKD was estimated using three definitions: (1): an estimated glomerular filtration rate (eGFR) <60 mL/min/1.73m^2^ or an eGFR ≥60 mL/min/1.73m^2^ with a UACR ≥2.5 mg/mmol for males and ≥3.5 mg/mmol for females (definition 1), (2) two consecutive eGFR measures <60 mL/min/1.73m^2^ at least 90 days apart or an eGFR ≥60 mL/min/1.73m^2^ with a UACR ≥2.5 mg/mmol for males and ≥3.5 mg/mmol for females (definition 2), and (3) two consecutive eGFR measures <60 mL/min/1.73m^2^ at least 90 days part and/or two consecutive UACR measures ≥2.5 mg/mmol for males and ≥3.5 mg/mmol for females at least 90 days apart (definition 3). Patient sociodemographic characteristics including comorbid conditions were assessed across the three definitions.

**Results:** The prevalence of CKD in the study cohort progressively increased over the 10-year study period, irrespective of the method used to define CKD. The annual prevalence of CKD varied across the three CKD definitions, with definition 1 resulting in the highest estimates. In 2020, the prevalence of CKD in the study cohort was 8.4% (n=123,988), 4.7% (n=69,110) and 3.1% (n=45,360) using definitions 1, 2 and 3, respectively. The number of patients with UACR measurements was low such that, among those identified as having CKD in 2020, only 3.8%, 3.2% and 1.5% respectively, had both eGFR and UACR measurements available in the corresponding year. Patients in whom both eGFR and UACR measurements were available mostly had moderate or high risk of CKD progression by local and international CKD guidelines (83.6%, 80.6% and 76.2%, respectively). Comorbid burden in patients with CKD was also frequently observed (coronary heart disease: 28.9%, type 2 diabetes: 38.5%, heart failure: 17.9%; using CKD definition 3).

**Conclusion:** In this large, nationally representative study, we observed an increasing trend in CKD prevalence in primary care settings in Australia. Most patients with CKD were at moderate to high risk of CKD progression with a significant comorbid burden including coronary heart disease and diabetes. These findings highlight the need for early detection and effective management to slow progression of CKD.

## INTRODUCTION

Chronic kidney disease (CKD), defined as impaired kidney function (estimated glomerular filtration [eGFR] <60 mL/min/1.73m^2^ that persists for ≥3 months), or the presence of other markers of kidney damage such as albuminuria (urine albumin-creatinine ratio [UACR] >3 mg/mmol),^1^ is a major global public health threat. CKD is tightly linked to other major chronic diseases, such as diabetes and cardiovascular disease, which collectively remain the leading causes of morbidity and pre-mature death worldwide. With increasing levels of diabetes^2^ and hypertension^3^ worldwide, leading causes of CKD, the burden of CKD is expected to increase in the foreseeable future and cost-effective means of monitoring are essential.

In Australia, population estimates of CKD prevalence suggest that approximately 11%^4^ of adults have CKD. However, there remains substantial variation in reported prevalence estimates (2.3% to 11%).^5–9^ Further, much of the national effort to establish CKD burden in Australia has focused on end-stage kidney disease (e.g. Australia and New Zealand Dialysis and Transplant Registry^10^ [ANZDATA]), while the epidemiology of earlier stages of CKD has been less well characterised. Emerging studies of CKD epidemiology in Australian primary care (the clinical setting in which the majority of patients with CKD are managed) indicate a significant burden of disease. However, many of these studies have several limitations including outdated estimates^6, 7^, restricted geographical coverage^5, 8^, variation in CKD definitions^5, 7–9^ (including the number of eGFR and/or UACR measurements considered) and limited information on the staging of CKD according to levels of kidney function and albuminuria^9^. These limitations restrict our ability to accurately determine the CKD burden in Australia.

A comprehensive and contemporary assessment of the prevalence of CKD is needed to better delineate the burden of CKD in Australia and inform health system policies for the care of this patient population. Using a large, nationally representative general practice data source in Australia, we conducted a community-based cohort study to determine the prevalence and staging of CKD in Australian primary care.

## METHODS

### Study design and source population

We conducted a retrospective cohort study using healthcare data from MedicineInsight, a national source of general practice data in Australia. Details of the MedicineInsight data source is provided elsewhere.^11^ Briefly, MedicineInsight data include de-identified longitudinal patient information, including sociodemographic and clinical characteristics (e.g. comorbid conditions, laboratory test results and other clinical measurements), and prescription medications on patients visiting general practices participating in MedicineInsight from all states and territories in Australia.

### Identification of the study cohort

The study cohort included all adults (≥18 years) with ≥1 visit to a general practice participating in the MedicineInsight program with ≥1 serum creatinine measurement (with or without a urine albumin-to-creatinine ratio [UACR] measurement) between 1 January 2011 and 31 December 2020 (n=2,720,529) across 392 general practices (Figure 1).

**Figure 1:**
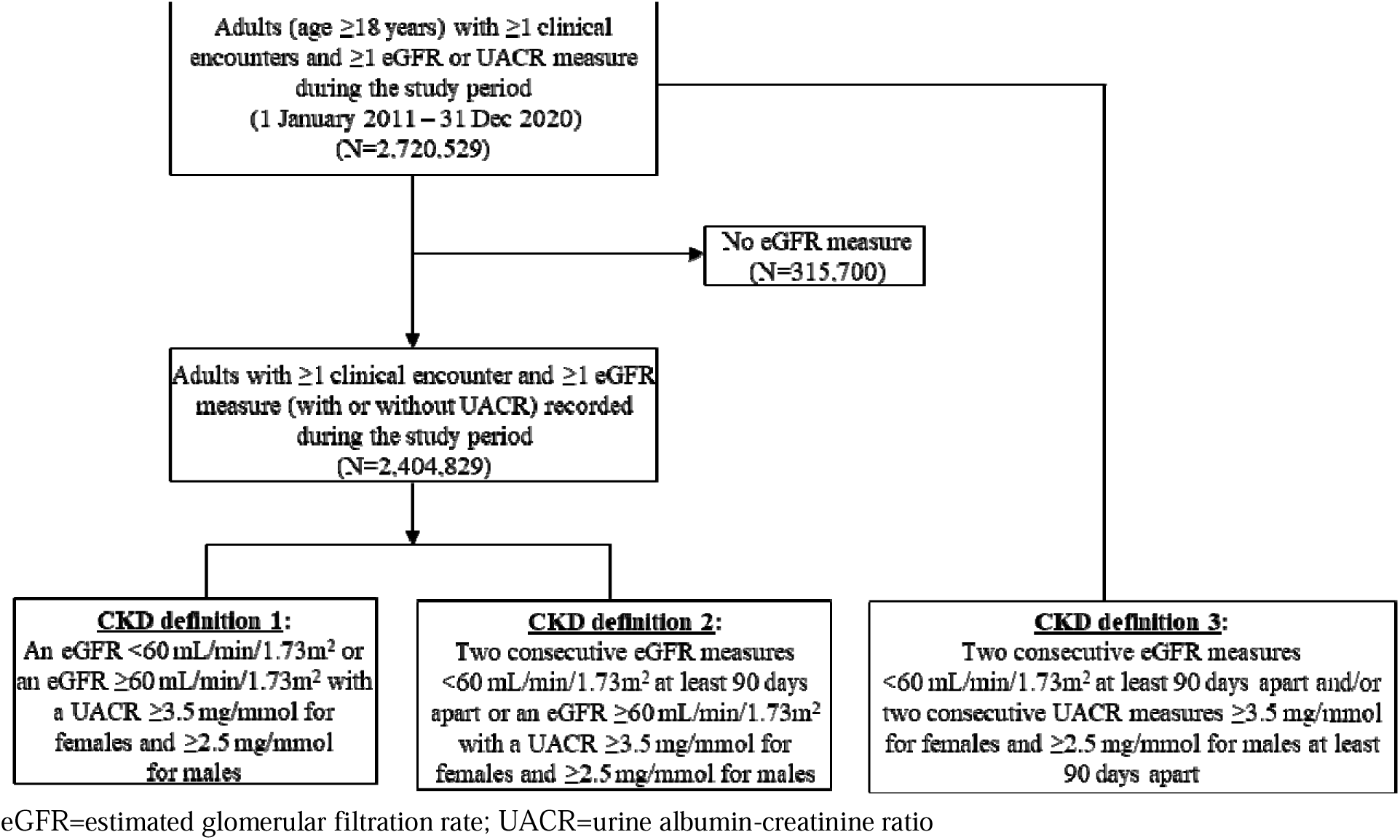
Identification of the study cohort.

### Prevalence of CKD

We estimated the prevalence of CKD in the study cohort using three methods based on local (Kidney Health Australia – Caring for Australian and New Zealanders with Kidney Impairment; KHA-CARI^12^; including sex-specific criteria for UACR), international (Kidney Disease: Improving Global Outcomes; KDIGO^1^) clinical practice guidelines for the management of patients with CKD and definitions used in prior published studies of CKD prevalence to enable between-study comparisons. CKD was defined in three ways (Supplementary table 1): (1) an estimated glomerular filtration rate (eGFR) <60 mL/min/1.73m^2^ (with or without albuminuria) or an eGFR ≥60 mL/min/1.73m^2^ with a UACR ≥2.5 mg/mmol for males and ≥3.5 mg/mmol for females (hereinafter referred to as “definition 1”), (2) two consecutive eGFR measures <60 mL/min/1.73m^2^ at least 90 days apart (with or without albuminuria) or an eGFR ≥60 mL/min/1.73m^2^ with a UACR ≥2.5 mg/mmol for males and ≥3.5 mg/mmol for females (“definition 2”), and (3) two consecutive eGFR measures <60 mL/min/1.73m^2^ at least 90 days part and/or two consecutive UACR measures ≥2.5 mg/mmol for males and ≥3.5 mg/mmol for females at least 90 days apart (“definition 3”). eGFR was derived using the Chronic Kidney Disease Epidemiology Collaboration (CKD-EPI) creatinine equation.^13^

Staging of CKD was based on eGFR (≥90, 60-89, 45-59, 30-44, 15-29 and <15 mL/min/1.73m^2^) and albuminuria (normal to mildly increased: <2.5 mg/mmol for males or <3.5 mg/mmol for females; moderately increased: 2.5-25 mg/mmol for males or 3.5-35 mg/mmol for females; severely increase: >25 mg/mmol for males or >35 mg/mmol for females) categories, as per the KHA-CARI and KDIGO clinical practice guidelines^1, 12^. Stages were defined as: Stage 1: eGFR ≥90 mL/min/1.73m^2^ and moderately or severely increased UACR; Stage 2: eGFR 60-89 mL/min/1.73m^2^ and moderately or severely increased UACR; Stage 3a: eGFR 45-59 mL/min/1.73m^2^; Stage 3b: eGFR 30-44 mL/min/1.73m^2^; Stage 4: eGFR 15-29 mL/min/1.73m^2^; Stage 5: eGFR <15 mL/min/1.73m^2^.

### Covariates

We obtained information on patient sociodemographic characteristics (sex, age, remoteness [major cities, inner regional, outer regional, remote and very remote], state [Australian Capital Territory, New South Wales, Northern Territory, Queensland, South Australia, Tasmania, Victoria and Western Australia], socioeconomic disadvantage (defined by Census variables based on neighbourhoods^14^: Index of Relative Socioeconomic Advantage and Disadvantage), smoking status and comorbid conditions (atrial fibrillation, cancer, coronary heart disease, chronic liver disease, chronic obstructive pulmonary disease, dementia, type 1 and 2 diabetes mellitus, heart failure, hypertension, peripheral vascular disease, rheumatoid arthritis, stroke or transient ischaemic attack) from the relevant data files of MedicineInsight.

### Statistical analysis

Continuous variables with approximately symmetric distributions are reported as means (with standard deviation [SD]), while those with skewed distributions are reported as median (with interquartile interval [IQI]). Categorical variables are presented as numbers and percentages. For each calendar year of the study period, the prevalence of CKD was calculated based on each patient’s first eligible eGFR and UACR measurement in the corresponding calendar year. Although information on the year of death is available in MedicineInsight, specific dates of death are not available. We therefore assigned “June 30” as the date for assessing deaths over the study period. The prevalence of CKD is reported as the number and percentage (as a proportion of the total number of eligible patients in each calendar year). A sensitivity analysis was performed in which CKD prevalence was calculated after removing sex-specific thresholds for the classification of albuminuria (normal to mildly increased: <3 mg/mmol; moderately increased: 3-30 mg/mmol; severely increased: >30 mg/mmol). All statistical analyses were performed with Stata version 17.0 (Stata, TX).

### Ethical approval and reporting standards

This study was approved by the MedicineInsight Data Governance Committee (2020-004) and Research Ethics Review Committee of the Sydney Local Health District, NSW, Australia (X21-0428, 2020/ETH00963). The study abides by the RECORD extension of the STROBE reporting guidelines^15^.

## RESULTS

### Cohort characteristics

Table 1 describes the patient characteristics of individuals identified as having CKD in 2020 according to the three CKD definitions assessed. In 2020, the mean age of patients identified based on definition 1 (n=123,988) was 74 years (SD 13) and 51.3% were female. Most individuals resided in major cities (54.5%), while 45.5% resided in regional or remote regions. Mean eGFR was 53.5 mL/min/1.73m^2^ (SD 17.2) and median UACR was 4.4 mg/mmol (IQI 1.6 – 11.6). Patients were distributed roughly evenly across the ten levels of increasing socioeconomic disadvantage defined. Coronary heart disease, type 2 diabetes and hypertension were present among 23.0%, 31.8% and 68.8% patients, respectively (Definition 1; Table 1, Supplementary figure 1).

**Table 1:** Characteristics of people with chronic kidney disease in MedicineInsight in 2020.

|  | No CKD* | Definition 1 | Definition 2 | Definition 3 |
| --- | --- | --- | --- | --- |
|  | N=1,345,845 | N=123,988 (8.4%) | N=69,110 (4.7%) | N=45,360 (3.1%) |
| **Sociodemographic information** |  |  |  |  |
| Sex; |  |  |  |  |
| Female | 787279 (58.5) | 63596 (51.3) | 32491 (47.0) | 22976 (50.7) |
| Male | 558566 (41.5) | 60392 (48.7) | 36619 (53.0) | 22384 (49.3) |
| Age (years); mean (SD) | 50.0 (18.2) | 74.3 (13.1) | 72.5 (14.0) | 76.7 (11.4) |
| Age category (years); |  |  |  |  |
| 18-29 | 214108 (15.9) | 782 (0.6) | 657 (1.0) | 124 (0.3) |
| 30-39 | 219968 (16.3) | 1852 (1.5) | 1520 (2.2) | 338 (0.7) |
| 40-49 | 226262 (16.8) | 3931 (3.2) | 3017 (4.4) | 809 (1.8) |
| 50-59 | 241966 (18.0) | 9046 (7.3) | 6137 (8.9) | 2185 (4.8) |
| 60-69 | 222265 (16.5) | 20239 (16.3) | 11939 (17.3) | 6191 (13.6) |
| 70-79 | 152521 (11.3) | 40236 (32.5) | 21732 (31.4) | 15370 (33.9) |
| ≥80 | 68755 (5.1) | 47902 (38.6) | 24108 (34.9) | 20343 (44.8) |
| Remoteness^; |  |  |  |  |
| Major cities | 817498 (61.0) | 67316 (54.5) | 37931 (55.1) | 24557 (54.3) |
| Inner regional | 348024 (26.0) | 38490 (31.2) | 21156 (30.7) | 14608 (32.3) |
| Outer regional | 157578 (11.8) | 16398 (13.3) | 9021 (13.1) | 5640 (12.5) |
| Remote | 12903 (1.0) | 1031 (0.8) | 590 (0.9) | 322 (0.7) |
| Very remote | 4296 (0.3) | 299 (0.2) | 178 (0.3) | 85 (0.2) |
| State; |  |  |  |  |
| Australian Capital Territory | 34561 (2.6) | 2167 (1.8) | 1190 (1.7) | 776 (1.7) |
| New South Wales | 444798 (33.1) | 45286 (36.5) | 25088 (36.3) | 17012 (37.5) |
| Northern Territory | 10055 (0.8) | 810 (0.7) | 446 (0.7) | 215 (0.5) |
| Queensland | 276943 (20.6) | 22485 (18.1) | 12325 (17.8) | 8692 (19.2) |
| South Australia | 28473 (2.1) | 2538 (2.1) | 1413 (2.0) | 997 (2.2) |
| Tasmania | 82063 (6.1) | 9982 (8.1) | 5956 (8.6) | 3661 (8.1) |
| Victoria | 304250 (22.6) | 27999 (22.6) | 15140 (21.9) | 9650 (21.3) |
| Western Australia | 164702 (12.2) | 12721 (10.3) | 7552 (10.9) | 4357 (9.6) |
| 2016 Index of Relative Socio-Economic Advantage and Disadvantage (IRSAD) decile; |  |  |  |  |
| 1 (most disadvantaged) | 103927 (7.8) | 12641 (10.2) | 7537 (10.9) | 4984 (11.0) |
| 2 | 120555 (9.0) | 14576 (11.8) | 8007 (11.6) | 5445 (12.0) |
| 3 | 93608 (7.0) | 10069 (8.2) | 5709 (8.3) | 3703 (8.2) |
| 4 | 161223 (12.0) | 17365 (14.1) | 9714 (14.1) | 6438 (14.2) |
| 5 | 152921 (11.4) | 15234 (12.3) | 8487 (12.3) | 5700 (12.6) |
| 6 | 145239 (10.8) | 12129 (9.8) | 6835 (9.9) | 4265 (9.4) |
| 7 | 122380 (9.1) | 9954 (8.1) | 5637 (8.2) | 3659 (8.1) |
| 8 | 147013 (11.0) | 9916 (8.0) | 5585 (8.1) | 3481 (7.7) |
| 9 | 159559 (11.9) | 12308 (10.0) | 6429 (9.3) | 4297 (9.5) |
| 10 (most advantaged) | 133886 (10.0) | 9344 (7.6) | 4937 (7.2) | 3241 (7.2) |
| Smoking status; |  |  |  |  |
| Smoker | 165167 (12.3) | 8677 (7.0) | 5518 (8.0) | 2419 (5.3) |
| Previous smoker | 291620 (21.7) | 43731 (35.3) | 25685 (37.2) | 17477 (38.5) |
| Non-smoker | 774613 (57.6) | 63788 (51.4) | 34519 (49.9) | 23283 (51.3) |
| Not recorded | 114445 (8.5) | 7792 (6.3) | 3388 (4.9) | 2181 (4.8) |
| **eGFR category (mL/min/1.73m<sup>2</sup>);** |  |  |  |  |
| ≥90 | 388944 (55.1) | 9482 (7.6) | 9482 (13.7) | 1614 (3.6) |
| 60-89 | 317042 (44.9) | 13199 (10.6) | 13199 (19.1) | 2313 (5.1) |
| 45-59 | - | 69025 (55.7) | 26222 (37.9) | 22945 (50.6) |
| 30-44 | - | 23853 (19.2) | 14478 (20.9) | 13261 (29.2) |
| 15-29 | - | 6884 (5.6) | 4759 (6.9) | 4344 (9.6) |
| <15 | - | 1545 (1.2) | 970 (1.4) | 883 (1.9) |
| eGFR (mL/min/1.73m<sup>2</sup>), mean (SD) | 84.5 (8.2) | 53.5 (17.2) | 56.6 (20.9) | 47.1 (15.7) |
| UACR (mg/mmol), median (IQI) | 0.8 (0.5 – 1.3) | 4.4 (1.6 – 11.6) | 5.8 (3.1 – 15.1) | 4.6 (1.4 – 15.6) |
| **Comorbid conditions; n (%)** |  |  |  |  |
| Atrial fibrillation | 37659 (2.8) | 20533 (16.6) | 11994 (17.4) | 9677 (21.3) |
| Cancer | 226335 (16.8) | 46943 (37.9) | 25878 (37.4) | 19520 (43.0) |
| Coronary heart disease | 64166 (4.8) | 28471 (23.0) | 17340 (25.1) | 13101 (28.9) |
| Chronic liver disease | 3796 (0.3) | 1104 (0.9) | 707 (1.0) | 522 (1.2) |
| Chronic obstructive pulmonary disease | 49790 (3.7) | 16328 (13.2) | 9392 (13.6) | 7046 (15.5) |
| Dementia | 6987 (0.5) | 3376 (2.7) | 1440 (2.1) | 1204 (2.7) |
| Diabetes mellitus |  |  |  |  |
| Type 1 | 8762 (0.7) | 3195 (2.6) | 2570 (3.7) | 1476 (3.3) |
| Type 2 | 79657 (5.9) | 39459 (31.8) | 29765 (43.1) | 17455 (38.5) |
| Heart failure | 15917 (1.2) | 15171 (12.2) | 9522 (13.8) | 8110 (17.9) |
| Hypertension | 317691 (23.6) | 85251 (68.8) | 49922 (72.2) | 34063 (75.1) |
| Peripheral vascular disease | 7392 (0.5) | 5138 (4.1) | 3313 (4.8) | 2539 (5.6) |
| Rheumatoid arthritis | 17258 (1.3) | 3870 (3.1) | 2317 (3.4) | 1848 (4.1) |
| Stroke or transient ischaemic attack | 30755 (2.3) | 13345 (10.8) | 7537 (10.9) | 5885 (13.0) |
| **Number of comorbidities#; n (%)** |  |  |  |  |
| 0 | 806154 (59.9) | 12583 (10.1) | 4999 (7.2) | 2393 (5.3) |
| 1-2 | 463371 (34.5) | 62988 (50.8) | 33377 (48.3) | 20052 (44.2) |
| ≥3 | 76320 (5.6) | 48417 (39.0) | 30734 (44.5) | 22915 (50.6) |

CKD=chronic kidney disease; \*Defined as people not categorised as having CKD by any of the 3 CKD definitions used; SD=standard deviation ^2016 Australian Statistical Geography Standard (ASGS) Remoteness Area; eGFR=estimated glomerular filtration rate; UACR=urine albumin-creatinine ratio; IQI=interquartile interval; #Based on the listed comorbid conditions

Compared to patients identified as having CKD based on definition 1, those with CKD based on definition 3 (n=69,110) were older (mean age of 76.7 vs, 74.3 years), had lower eGFR (mean eGFR 47.1 vs 53.5 mL/min/1.73m^2^) and greater comorbidity (Table 1).

### Prevalence of CKD

The derivation of the cohorts from the MedicineInsight data using the three definitions is outlined in Figure 1. In 2020, 123,988 (8.4%), 69,110 (4.7%) and 45,360 (3.1%) people were identified as having CKD in the study cohort based on definition 1, 2 and 3, respectively. During the 10-year study period (2011–2020), the prevalence of CKD progressively increased, irrespective of the definition used (Figure 2). However, the annual prevalence of CKD varied across the three CKD definitions, with stricter definitions (definitions 2 and 3) resulting in lower estimates. The number of patients with UACR measurements was low such that, of the individuals identified as having CKD in 2020, only 3.8%, 3.2% and 1.5% respectively, had both eGFR and UACR measurements available in the corresponding year to enable stratification of CKD according to both markers (Figure 3). Among patients in whom both eGFR and UACR measurements were available, 83.6%, 80.6% and 76.2%, respectively, had moderate or high risk of CKD progression (“yellow” and “orange” risk groups in Figure 3; Supplementary figure 2). The overall prevalence of CKD was highest in patients aged ≥80 years (ranging from 16.8% to 41.8% across CKD definitions) and higher among males than females across all age categories; Supplementary table 2). Prevalence estimates were similar when sex-specific UACR thresholds were removed (Supplementary figure 3, Supplementary table 3).

**Figure 2:**
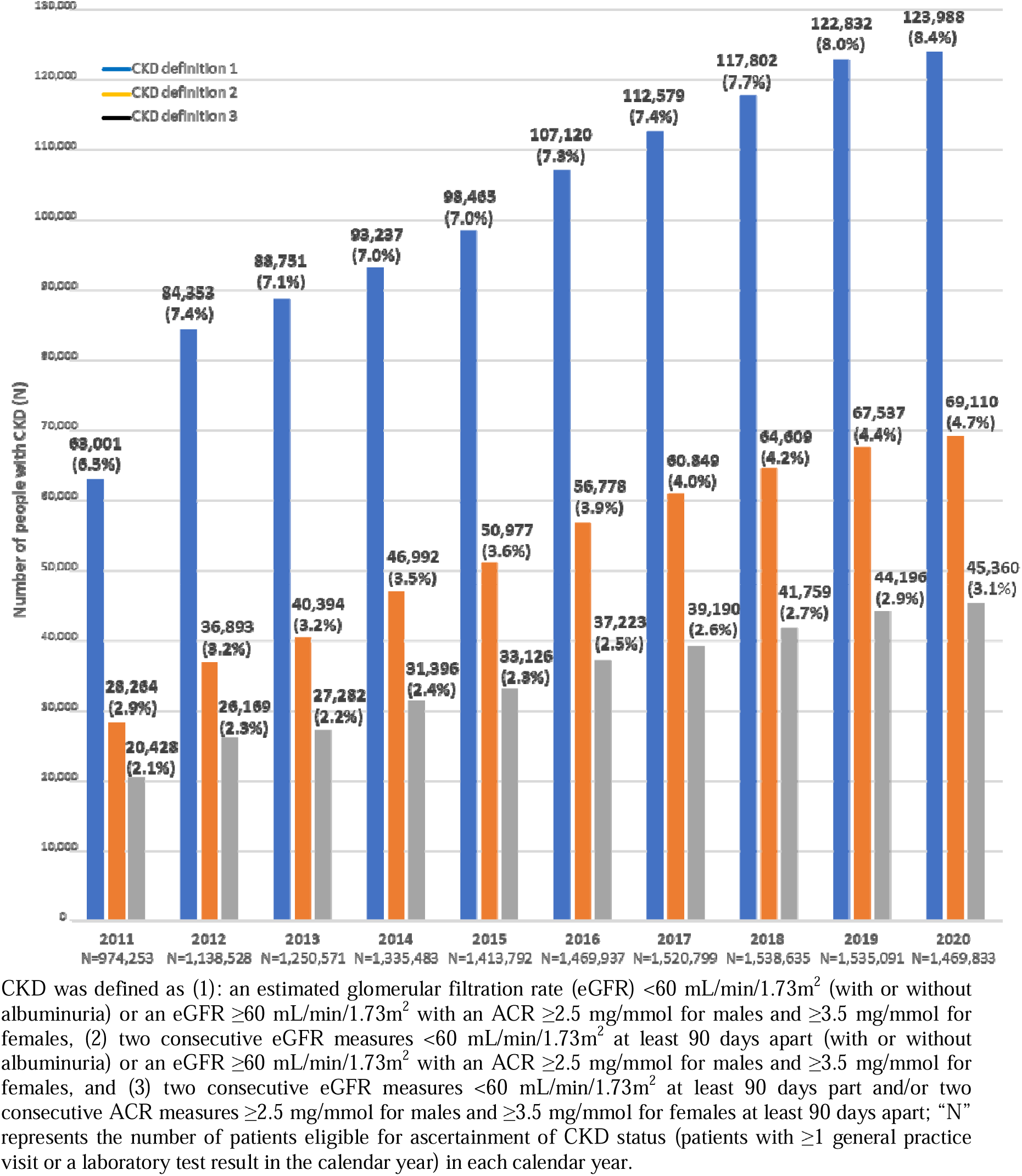
Annual prevalence of chronic kidney disease in MedicineInsight (2011-2020)

**Figure 3:**
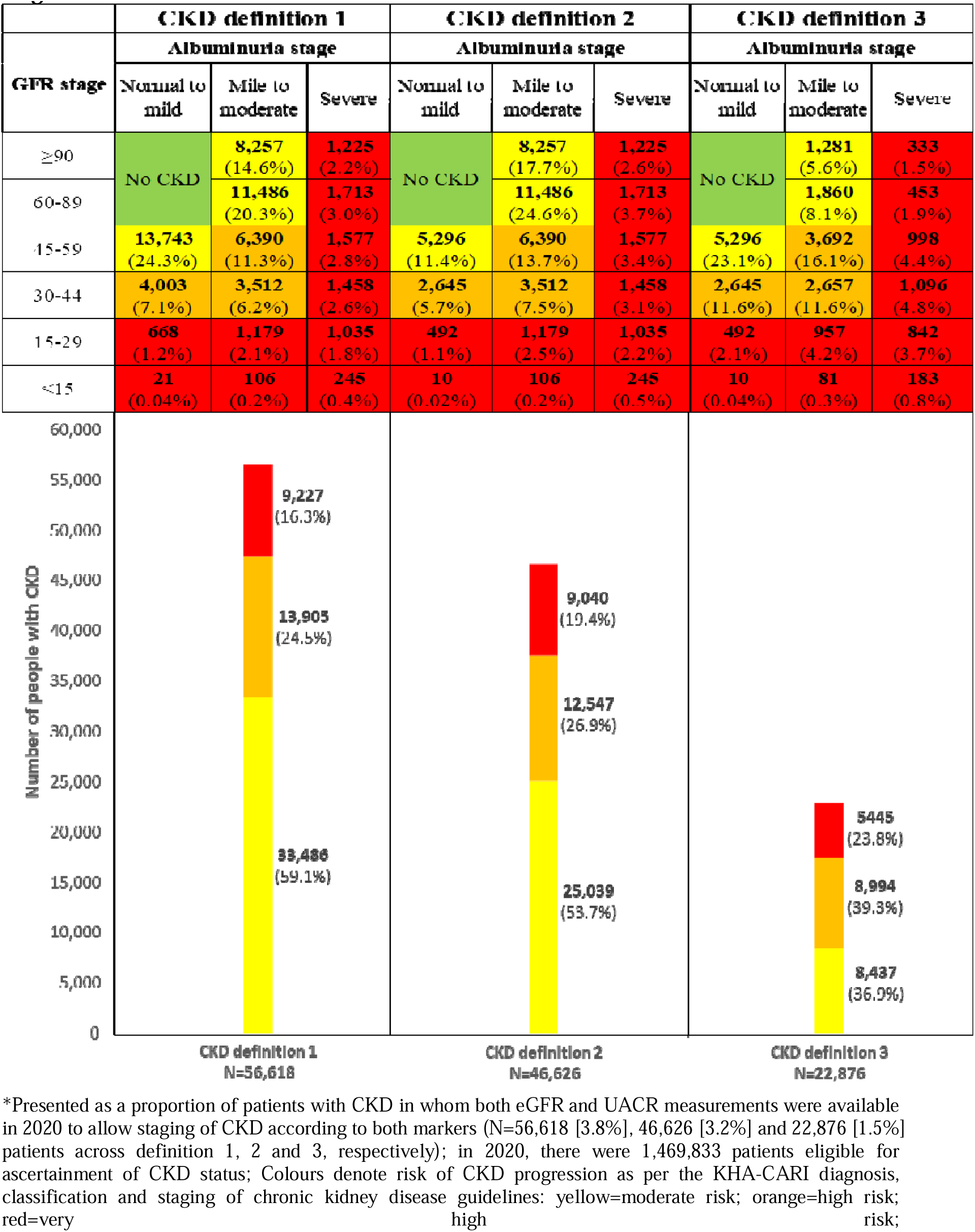
Chronic kidney disease prevalence in 2020* in MedicineInsight by KDIGO stage.

## DISCUSSION

Based on a large, nationally representative primary care data source, this community-based study shows that the prevalence of CKD has been increasing over the last decade in Australia, irrespective of the definition of CKD. In 2020, the prevalence of CKD ranged from 3.1% to 8.4% depending upon the rigidity of the CKD definition used (i.e. the number of eGFR and/or UACR measurements used to define CKD). UACR testing rates are low and among those in whom risk stratification according to KDIGO CKD guidelines was possible, most patients with CKD were at moderate to high risk of CKD progression. Comorbid burden in patients with CKD was also frequently observed. Coronary heart disease was present in 23.0%-28.9% of patients, while type 2 diabetes and heart failure were present in 31.8%-43.1% and 12.2%-17.9%, respectively. Taken together, these results highlight the increasing burden of CKD in Australia and the need to effectively implement management strategies aimed at improving outcomes in this patient population.

Prior estimates from ANZDATA indicate increasing prevalence of kidney replacement therapy between 2010 to 2020 in Australia.^10^ Our study extends this finding to the broader spectrum of CKD at a national level, showing a similar increasing trajectory in the burden of CKD among those with early or moderate stages of CKD. Our results also suggest that most patients with CKD managed in primary care are at moderate to high risk of progression with a high burden of comorbidity – over 1 in 4 patients with CKD had coronary heart disease, while over 1 in 3 had type 2 diabetes. These results align with recent international data^16, 17^ including a study of 7 other countries showing that 69% of patients with CKD in those countries have CKD stage 3-4, with significant comorbid burden^17^. Such findings emphasise the importance of prevention strategies that delay disease progression and prevent onset of adverse outcomes, including cardiovascular disease. Effective treatments have recently emerged including sodium-glucose co-transporter 2 inhibitors^18^, building on the beneficial effects observed with long-standing agents such as angiotensin-receptor blockade.^1^ To achieve optimal implementation of these treatment options in routine care, a better understanding of current patterns of CKD care across different patient groups and settings is needed. This will help to monitor processes of care, compare concordance of clinical practice with guideline recommendations and identify areas for improvement in this high-risk group – a patient population in whom the greatest magnitude of benefit from effective treatments may be seen. While such studies in Australia have been limited, the increasing availability of large, routinely collected health data sources such as MedicineInsight will help to enable this work. Results from our current study can serve as a basis upon which to develop such efforts.

Previous studies on CKD prevalence in Australia have reported estimates ranging between 2.3% and 11.5%.^4–9^ The prevalence of CKD in 2020 in the present study (up to 8.4%) lies within this range but varies according to the method used to define CKD (single vs. multiple assessments of kidney function and albuminuria). Indeed, much of the heterogeneity in reported CKD prevalence estimates to date may be attributed to differences in methods for defining CKD. For example, Radford et al^9^, also using data from MedicineInsight (but based on an earlier cross-section of the data; 2014-2016), reported that CKD prevalence in 2016 was 5.2% using a single measurement-based approach and 4.1% based on two eGFR and/or UACR measurements. Our study, based on a more contemporaneous and larger cohort of patients, confirms this, showing variable CKD prevalence according to the number of eGFR and UACR measurements used. Of note, there were some differences in the estimates of CKD prevalence between the two studies (e.g., CKD prevalence of 2.5% in 2016 based on definition 3 in the present study compared to 4.1% in the study by Radford et al) which may be attributable to differences in cohort size and inclusion criteria (e.g., increased patient cohort size arising from greater participation of general practices in MedicineInsight). Nevertheless, these results highlight the importance of considering chronicity of kidney dysfunction when measuring CKD burden, particularly in the surveillance of disease progression and assessment of patient care patterns to inform quality improvement strategies.

This study has several strengths. Our study was based a large, community-based nationally representative data source inclusive of various geographic regions (including urban and remote; and all states and territories of Australia) which expand the generalisability of its findings. Assessment of the prevalence of CKD and its severity was based on a longitudinal analysis of data encompassing a 10-year period reflecting contemporary routine primary care settings and based on rigorous approaches to defining CKD. However, our study has some limitations that should be considered. While the data source captures a large national cohort of patients who visited a general practice, we do not have information on patients who visited general practices not participating in the MedicineInsight program; although, regular MedicineInsight patients are generally representative of the Australian population in terms of age and sex.^11^ Further, there are other factors that may potentially impact the estimation of CKD prevalence that warrant consideration. A substantial proportion of patients did not have a measure of UACR which may have led to an underestimation of the prevalence of individuals with earlier stages of CKD. Given that the “denominator” in estimating the prevalence of CKD was those who had a record of ever having an eGFR or UACR measurement during the study period, it is possible that this may have contributed to an overestimation of CKD. Nevertheless, overall results were generally aligned with those derived from comparable data sources in previous large studies across numerous countries^17, 19, 20^ which support the robustness of our study findings.

In summary, in this large, nationally representative community-based cohort study, we observed an increasing trend in CKD prevalence in primary care settings in Australia. Most patients with CKD were at moderate to high risk of CKD progression with comorbid burden including coronary heart disease and diabetes frequently observed. These findings highlight the importance of prevention, early detection and effective management of CKD including the need to monitor processes of care, identify potential areas for improvement and develop new interventions to improve outcomes in this high-risk group.

## Data Availability

The current study is based on data from MedicineInsight, a national general practice data source developed by NPS MedicineWise and managed by the Australian Commission on Safety and Quality in Health Care. All relevant data are within the manuscript and its supplementary appendix.

## AUTHOR CONTRIBUTIONS

MJ, JW, BLN, SK, SVB, MG and PER contributed to the concept and rationale for the study and interpretation of the results. KN and MJ were responsible for acquisition of the data extract from MedicineInsight. MJ, JW and PR developed the study protocol and oversaw the implementation of the study analytical plan. MJ drafted the initial manuscript. All authors contributed to the design of the study, interpretation of the data and critical revision of the manuscript.

## ACKNOWLEDGEMENTS

This study is based on data from MedicineInsight (project 2020-004), a national general practice data program developed by NPS MedicineWise and managed by the Australian Commission on Safety and Quality in Health Care. MedicineInsight extracts and collates longitudinal, de-identified, patient health data from the clinical information systems of consenting general practices across Australia. This study was supported by an unrestricted research grant from Boehringer Ingelheim. MJ is supported by the Scientia Program at the Faculty of Medicine and Health, UNSW Sydney, Australia.

## SUPPLEMENTARY APPENDIX

**Supplementary table 1:** Definitions of chronic kidney disease used in the current study.

| CKD definition 1 |  |  |
| --- | --- | --- |
| GFR stage<br>(mL/min/1.73m<sup>2</sup>) | Albuminuria stage |  |
|  | Normal to mild* | Mild to severe^ |
| ≥60 | No CKD | CKD (1, 1)<sup>#</sup> |
| <60 | CKD (1, 0) | CKD (1, 0-1) |
| CKD definition 2 |  |  |
| GFR stage<br>(mL/min/1.73m<sup>2</sup>) | Albuminuria stage |  |
|  | Normal to mild | Mild to severe |
| ≥60 | No CKD | CKD (1, 1) |
| <60 | CKD (2, 0) | CKD (2, 0-1) |
| CKD definition 3 |  |  |
| GFR stage<br>(mL/min/1.73m<sup>2</sup>) | Albuminuria stage |  |
|  | Normal to mild | Mild to severe |
| ≥60 | No CKD | CKD (0, 2) |
| <60 | CKD (2, 0) | CKD (2, 2) |

CKD=chronic kidney disease; GFR=glomerular filtration rate; \*Defined as urinary albumin-creatinine ratio (UACR) ≤2.5 mg/mmol for males and ≤3.5 mg/mmol for females; ^Defined as ACR >2.5 mg/mmol for males and >3.5 mg/mmol for females; #numbers in parentheses indicate the number of estimated GFR and UACR measurements, respectively, used to define CKD status

**Supplementary table 2:**
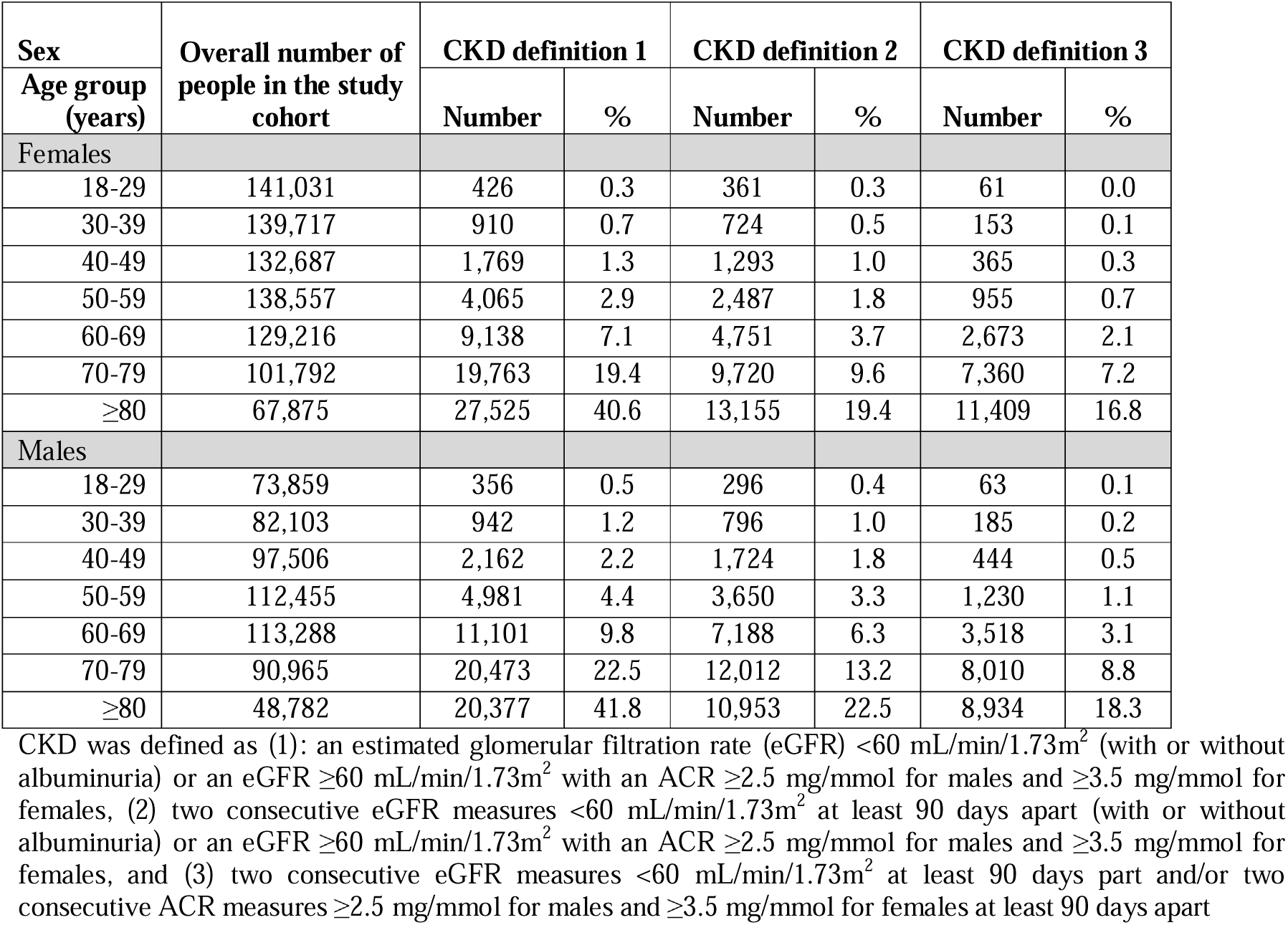
Prevalence of chronic kidney disease in MedicineInsight in 2020 by sex and age group.

**Supplementary table 3:** Prevalence of chronic kidney disease in MedicineInsight in 2020 by sex and age group after removing sex-specific thresholds for UACR.

| Sex | Overall number of people in the study cohort | CKD definition 1 |  | CKD definition 2 |  | CKD definition 3 |  |
| --- | --- | --- | --- | --- | --- | --- | --- |
| Age group (years) |  | Number | % | Number | % | Number | % |
| Females |  |  |  |  |  |  |  |
| 18-29 | 141,031 | 472 | 0.3 | 407 | 0.3 | 62 | 0.1 |
| 30-39 | 139,717 | 993 | 0.7 | 809 | 0.6 | 162 | 0.1 |
| 40-49 | 132,687 | 1,934 | 1.5 | 1,459 | 1.1 | 387 | 0.3 |
| 50-59 | 138,557 | 4,317 | 3.1 | 2,750 | 2.0 | 994 | 0.7 |
| 60-69 | 129,216 | 9,516 | 7.4 | 5,169 | 4.0 | 2,710 | 2.1 |
| 70-79 | 101,792 | 20,110 | 19.8 | 10,159 | 10.0 | 7,425 | 7.3 |
| ≥80 | 67,875 | 27,662 | 40.8 | 13,398 | 19.7 | 11,437 | 16.9 |
| Males |  |  |  |  |  |  |  |
| 18-29 | 73,859 | 327 | 0.4 | 266 | 0.4 | 61 | 0.1 |
| 30-39 | 82,103 | 864 | 1.1 | 718 | 0.9 | 173 | 0.2 |
| 40-49 | 97,506 | 1,982 | 2.0 | 1,537 | 1.6 | 432 | 0.4 |
| 50-59 | 112,455 | 4,662 | 4.2 | 3,315 | 3.0 | 1,169 | 1.0 |
| 60-69 | 113,288 | 10,651 | 9.4 | 6,685 | 5.9 | 3,407 | 3.0 |
| 70-79 | 90,965 | 20,046 | 22.0 | 11,462 | 12.6 | 7,884 | 8.7 |
| ≥80 | 48,782 | 20,248 | 41.5 | 10,732 | 22.0 | 8,899 | 18.2 |

CKD was defined as (1): an estimated glomerular filtration rate (eGFR) $<60 \text{ mL/min/1.73m}^2$ (with or without albuminuria) or an eGFR $\geq 60 \text{ mL/min/1.73m}^2$ with an ACR $\geq 2.5 \text{ mg/mmol}$ for males and $\geq 3.5 \text{ mg/mmol}$ for females, (2) two consecutive eGFR measures $<60 \text{ mL/min/1.73m}^2$ at least 90 days apart (with or without albuminuria) or an eGFR $\geq 60 \text{ mL/min/1.73m}^2$ with an ACR $\geq 2.5 \text{ mg/mmol}$ for males and $\geq 3.5 \text{ mg/mmol}$ for females, and (3) two consecutive eGFR measures $<60 \text{ mL/min/1.73m}^2$ at least 90 days part and/or two consecutive ACR measures $\geq 2.5 \text{ mg/mmol}$ for males and $\geq 3.5 \text{ mg/mmol}$ for females at least 90 days apart

**Supplementary figure 1:**
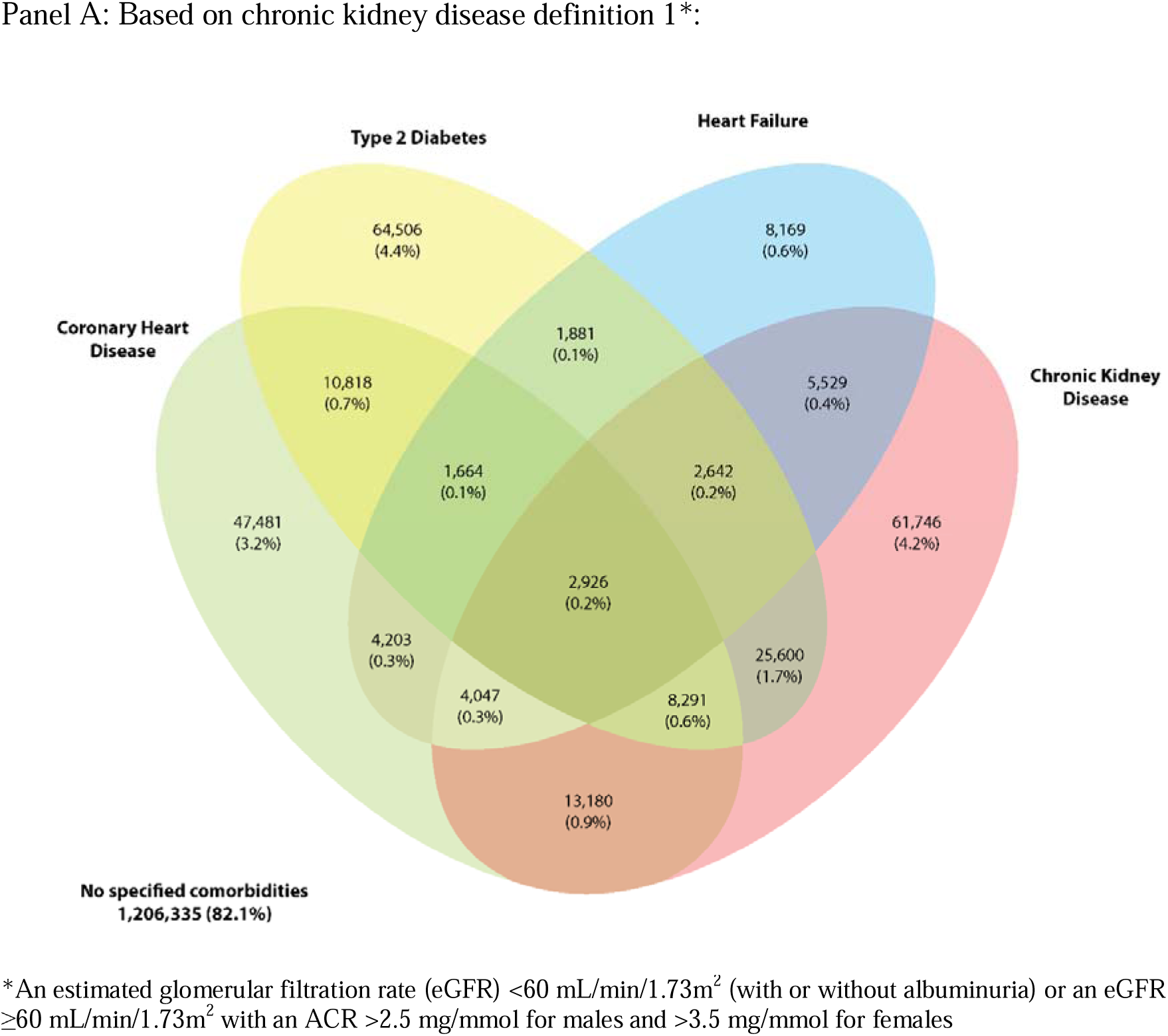
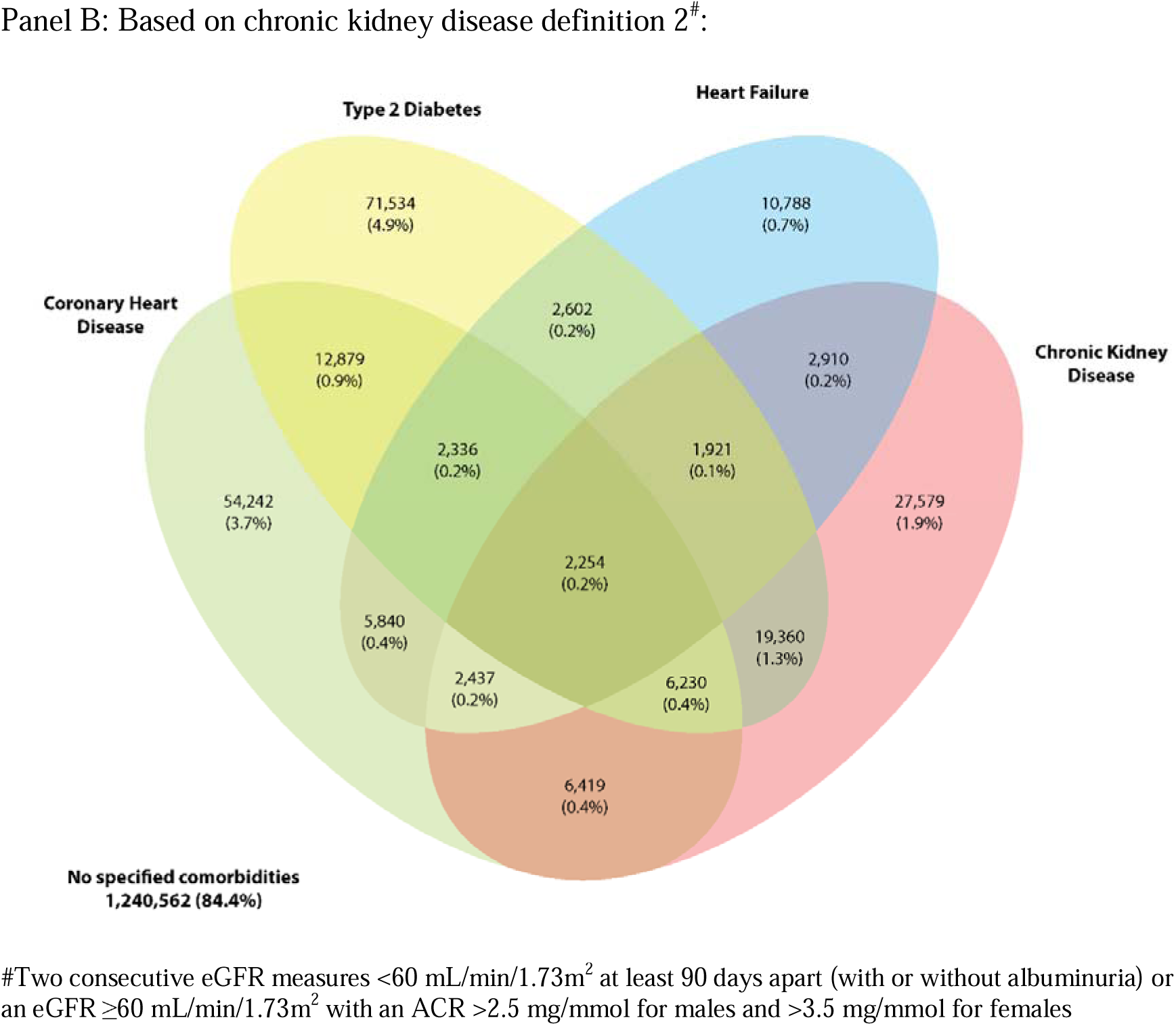
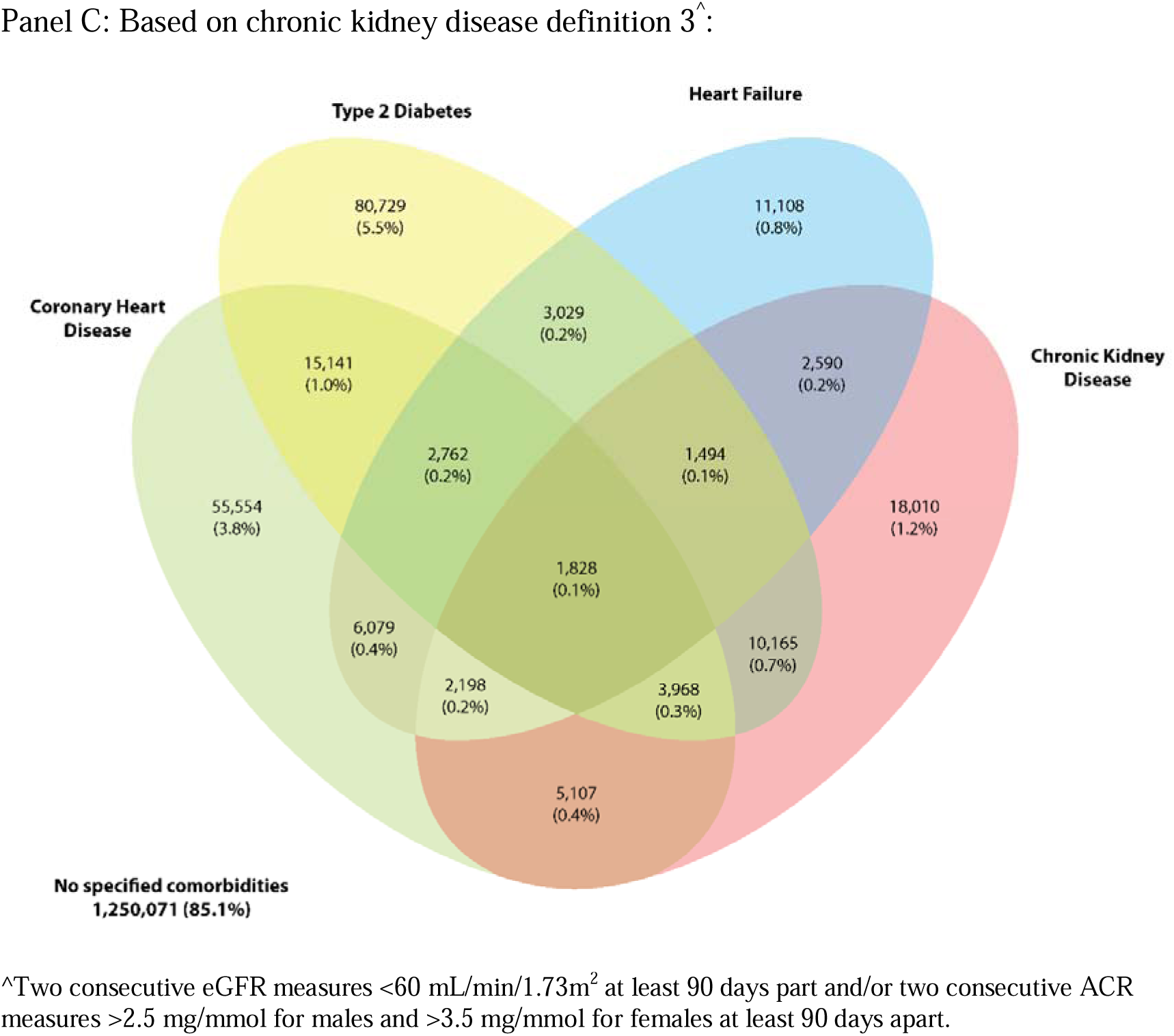
Venn diagram showing the relationship between chronic kidney disease and key comorbidities among patients in MedicineInsight in 2020.

**Supplementary figure 2:**
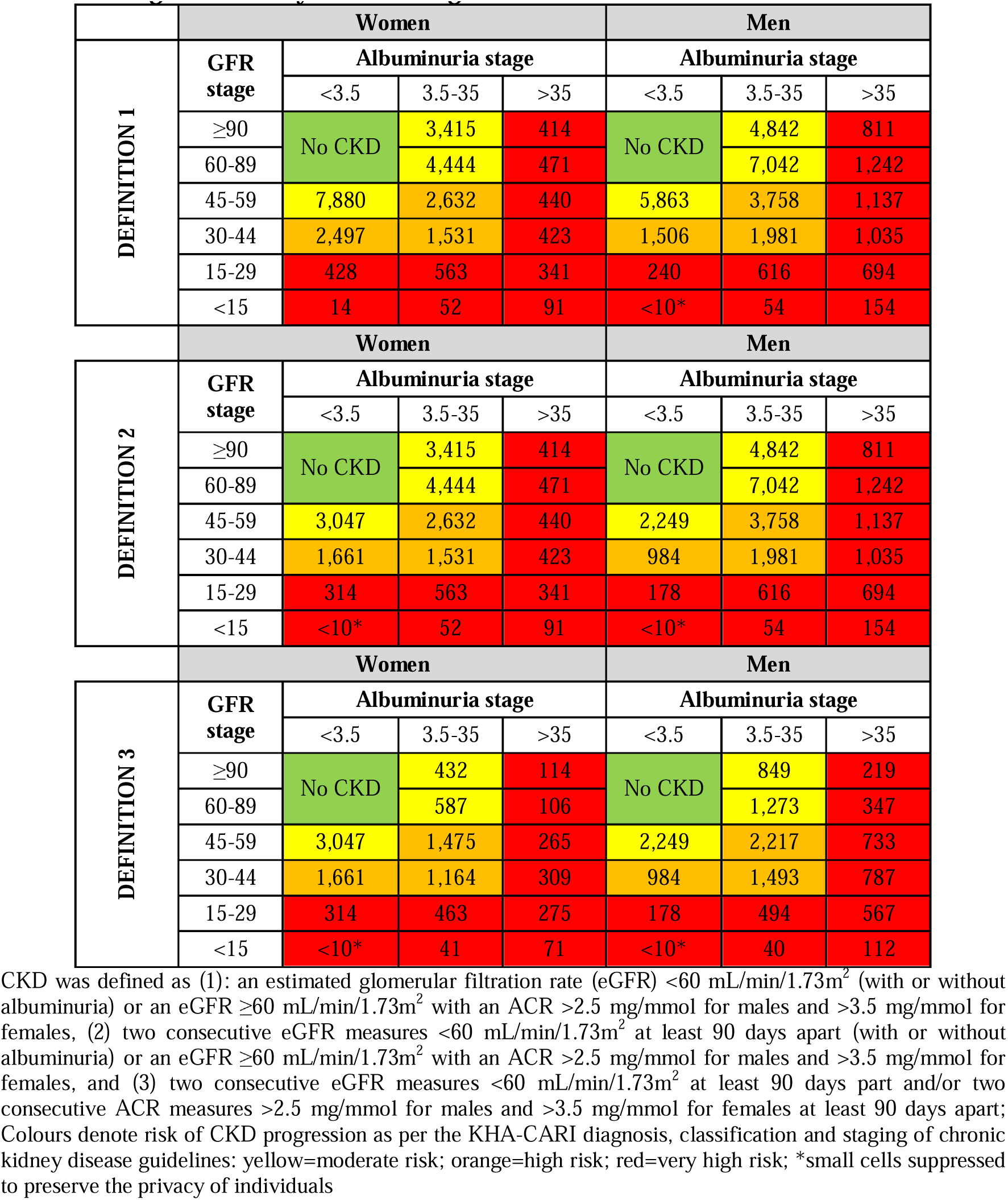
Sex-specific chronic kidney disease prevalence in MedicineInsight in 2020 by KDIGO stage.

**Supplementary figure 3:**
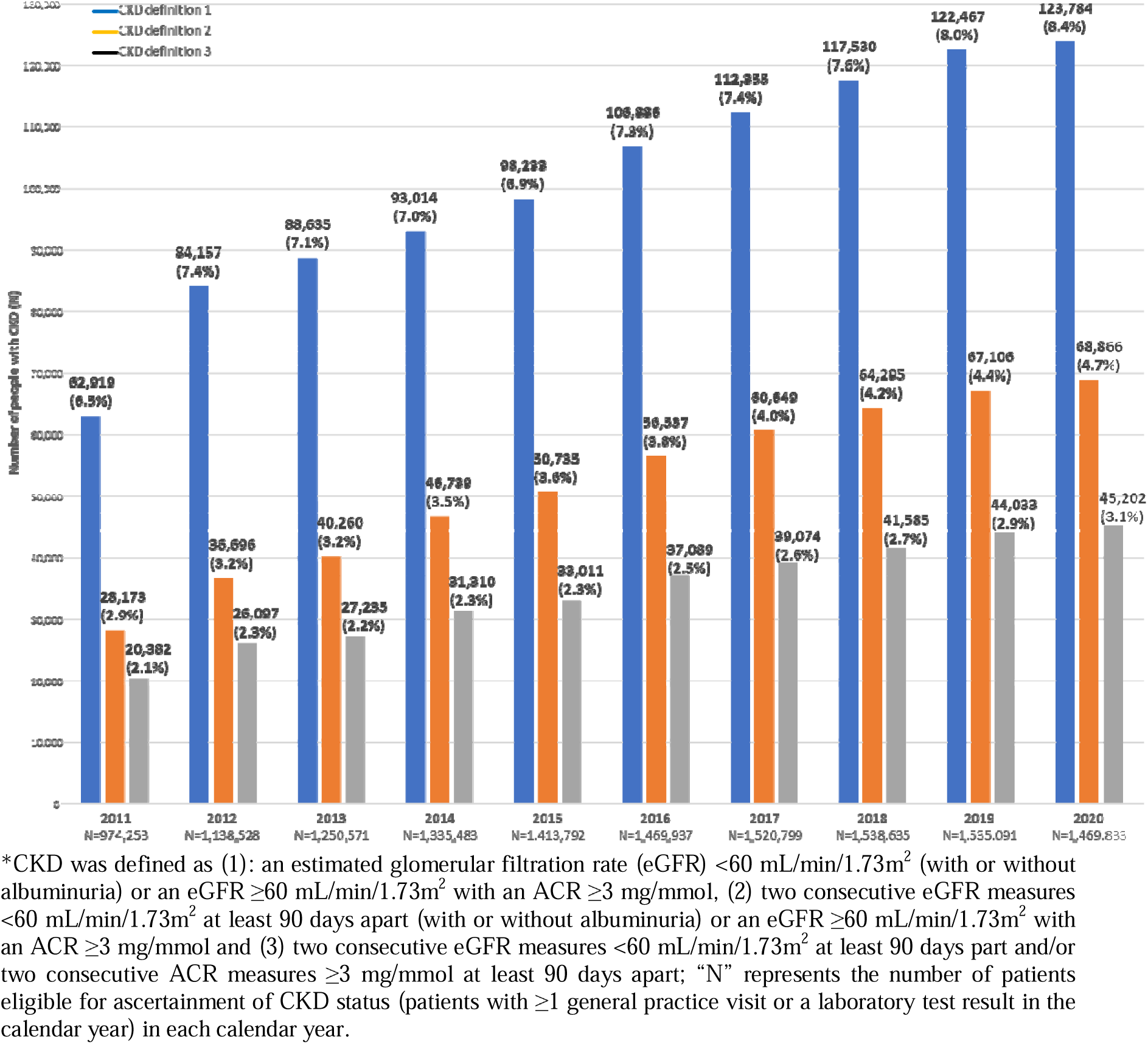
Annual prevalence of chronic kidney disease in MedicineInsight (2011-2020) after removing sex-specific thresholds for UACR*.

